# Automated brain morphometric biomarkers from MRI at term predict motor development in very preterm infants

**DOI:** 10.1101/2020.08.03.20167528

**Authors:** Julia E. Kline, Venkata Sita Priyanka Illapani, Lili He, Nehal A. Parikh

**Author notes:** Corresponding author’s contact information: Nehal A. Parikh, DO, MS, Professor of Pediatrics, Cincinnati Children’s Hospital, 3333 Burnet Ave, MLC 7009, Cincinnati, OH 45229, (Business).

## Abstract

Very preterm infants are at high risk for motor impairments. Early intervention can improve outcomes in this cohort, but it would be most effective if clinicians could accurately identify the highest risk infants early. A number of biomarkers for motor development exist, but currently none are sufficiently accurate for early risk-stratification. We prospectively enrolled very preterm (gestational age ≤31 weeks) infants from four level-III NICUs. Structural brain MRI was performed at term-equivalent age. We used a well-established pipeline to automatically derive brain volumetrics and cortical morphometrics, cortical surface area, sulcal depth, gyrification index, and inner cortical curvature, from structural MRI. We related these objective measures to Bayley-III motor scores (overall, gross, and fine) at two-years corrected age. Lasso regression identified the three best predictive biomarkers for each motor scale from our initial feature set. In multivariable regression, we assessed the independent value of these brain biomarkers, over-and-above known predictors of motor development, to predict motor scores. 75 very preterm infants had high-quality T2-weighted MRI and completed Bayley-III motor testing. All three motor scores were positively associated with regional cortical surface area and subcortical volumes and negatively associated with cortical curvature throughout the majority of brain regions. In multivariable regression modeling, thalamic volume, curvature of the temporal lobe, and curvature of the insula were significant predictors of overall motor development on the Bayley-III, independent of known predictors. Objective brain morphometric biomarkers at term show promise in predicting motor development in very preterm infants.

## 1. Introduction

Globally, slightly more than 10% of pregnancies result in preterm birth^1^. Very preterm infants (gestational age (GA) ≤ 31 weeks) are at greatly enhanced risk for cerebral palsy (CP) and also for less severe motor impairments, which, depending on GA, affect around 40% of preterm-born children^2,3^. Such impairments are often not diagnosed until at least two years of age, which means that clinicians and therapists miss a critical window for intervention, during which neuroplasticity is maximal. Given the proven efficacy of early intervention to enhance motor outcome^4^, a number of early motor biomarkers have been proposed^5^, but many require independent validation.

Of the possible structural brain biomarkers available at term-equivalent age (TEA), abnormality or injury on early cranial ultrasound or structural magnetic resonance imaging (MRI) have been widely used^6–8^, however neither is sufficient for accurate prediction. As such, researchers have turned to other structural brain features beyond overt injury. Brain volumes are consistently decreased in extremely-low-birth-weight or preterm cohorts^9–14^, and there is evidence that these volumetric changes prognosticate motor ability^13,15–21^. Preterm infants also exhibit altered cortical morphometrics, including inner cortical curvature^22,23^, surface area^14,22–28^, gyrification^14,23,28–30^, and sulcal depth^14,23,28,29^ at term. We previously identified a strong independent association of inner cortical curvature and cortical surface area with two-year cognitive and language outcomes in our very preterm cohort^31^, suggesting that these biomarkers may aid in the prediction of other early neurodevelopmental outcomes. An association between cortical surface area and motor ability has been previously established in preterm children and adults^9,32^. However, to our knowledge, no prior study has assessed the individual value of inner cortical curvature, sulcal depth, or gyrification index to prognosticate motor ability in preterm infants. A classifier trained on combined cortical morphometrics and tissue volumetrics from structural MRI at TEA had good success predicting abnormal two-year motor function in a preterm cohort^20^. However, neither the individual predictive power of each cortical morphometric nor the regional significance of the volumes were explored.

We aimed to independently validate and extend current knowledge about the predictive value of structural MRI biomarkers for motor outcome in very preterm infants. We examined the relationships of whole-brain and regional cortical morphometrics and volumetrics to overall, gross, and fine motor aptitude at two years of age. We then used methods from machine learning to identify the top predictive biomarkers for each motor outcome. We assessed their combined predictive ability in multivariable regression, corrected for known confounders of motor development. Our population-based cohort, ability to derive objective morphometric and volumetric features using an automated pipeline, and higher MRI field-strength (3T) than previous studies should allow us to pinpoint the most predictive morphometric MRI biomarkers of motor development available at TEA, which will lead to improved predictive models that facilitate earlier intervention in this high-risk cohort.

## 2. Materials and methods

### 2.1 Participants

Between December 2014 and April 2016, we prospectively enrolled a consecutive cohort of 110 very preterm infants (GA ≤ 31 weeks) from four regional, level-III neonatal intensive care units (NICUs) – Nationwide Children’s Hospital, Ohio State University Medical Center, Riverside Methodist Hospital, and Mount Carmel St. Ann’s Hospital. Babies were excluded if they had cyanotic heart disease or congenital or chromosomal anomalies of their central nervous system. Infants that remained hospitalized in the NICUs of Ohio State, Riverside, or St. Ann’s beyond 44 weeks postmenstrual age (PMA) were also excluded (N=7). This study was approved by Nationwide Children’s Hospital Institutional Review Board. A parent or guardian of each infant gave written informed consent before study enrollment.

### 2.2 MRI data acquisition

We acquired all MRI data at Nationwide Children’s Hospital using a 3T Siemens Skyra scanner and a 32-channel phased array head coil. Subjects were imaged at a mean (SD) PMA of 40.4 (0.6) weeks. A skilled neonatal nurse and neonatologist attended all scans. Each infant was fed 30 minutes prior to the scan and was fitted with silicone earplugs (Instaputty, E.A.R. Inc, Boulder, CO) and swaddled in a blanket and a vacuum immobilization device (MedVac, CFI Medical Solutions, Fenton, MI) to promote natural sleep. We used the following acquisition parameters: axial T2-weighted MRI: echo time 147 ms, repetition time 9500 ms, flip angle 150°, voxel dimensions 0.93×0.93×1.0 mm^3^, scan time 4:09 min; 3-dimensional magnetization-prepared rapid gradient-echo T1-weighted MRI: echo time 2.9 ms, repetition time 2270 ms, echo spacing time 8.5 ms, inversion time 1750 ms, flip angle 13°, voxel dimensions 1.0×1.0×1.0 mm^3^, scan time 3:32 min; axial susceptibility-weighted MRI : echo time 20 ms, repetition time 27 ms, flip angle 15°, voxel dimensions 0.7×0.7×1.6 mm^3^, scan time 3:11 min.

### 2.3 Motor outcomes

Our primary outcome of interest was the overall motor score from the Bayley Scales of Infant and Toddler Development–Third Edition (Bayley-III)^33^ standardized examination. Our secondary outcomes of interest were the gross and fine motor subscale scores. The Bayley-III overall motor score is a scaled composite of the gross motor and fine motor subtest scores, ranging from 40 to 160, with a mean (SD) of 100 (15). The entire cohort was scheduled for Bayley-III testing between 22 and 26 months corrected age. Four infants did not return during their appointment window, and were instead tested between 33- and 36-months corrected age. These four subjects were nonetheless included in the analysis, as the Bayley-III motor scores obtained in this later time window would be equally accurate or even more accurate than earlier scores.

### 2.4 MRI data processing

We used the automated, open-source pipeline from the developing Human Connectome Project (dHCP)^34^ to process all T2-weighted MRI scans. The dHCP pipeline performs tissue segmentation and volume estimation for each main tissue type and all cortical and subcortical structures. It also calculates the following metrics for each cortical region of the dHCP’s neonatal atlas: surface area, sulcal depth, gyrification index, inner cortical curvature, and cortical thickness, 510 metrics in all. We visually inspected each segmented scan and rejected any scans showing tissue identification errors, improper segmentation, or artifacts. Very preterm infants with moderate or severe ventriculomegaly were also excluded, as large ventricles were often inaccurately identified as white matter by the pipeline.

### 2.5 Statistical analysis

Because the dHCP generates a very large number of regional and merged morphometrics and volumetrics, variable reduction in several stages was necessary to find the most highly-correlated predictors and reduce the likelihood of type 1 errors. **Step 1) Initial feature selection:** as our initial cortical morphometric feature set, we chose four features that are known to be altered in prematurity^23^, surface area, gyrification index, sulcal depth, and inner cortical curvature, for the whole brain and bilaterally for all four brain lobes. We also included curvature and surface area for the insula bilaterally. Previously, we had found cortical thickness from the dHCP to not be correlated with outcomes from our cohort^31^, so we did not include thickness in the current analysis. As our initial set of volumetrics, we chose overall brain volume, bilateral volume of all four brain lobes and the insula, and volume of the main tissue types: cortical grey matter, white matter, deep grey matter, corpus callosum, CSF, and ventricles. We also included volumes for the main subcortical structures: the amygdala, hippocampus, caudate nucleus, lentiform nucleus, and subthalamic nucleus bilaterally and the corpus collosum, thalamus, brainstem and cerebellum unilaterally. **Step 2) Correlation with outcome:** From the 72 initial features described in step 1, we performed partial-correlation analysis, corrected for PMA at MRI scan, to identify variables that were significantly associated with each of the Bayley-III motor scales. We also accounted for false discovery rate in this step, using Benjamini Hochberg correction with a significance level of p<0.05. **Step 3) Iterative lasso regression for feature selection:** For each motor scale, we used least absolute shrinkage and selection operator (Lasso) regression^35^ to select the top features from the subset identified in step two. Lasso regression is a form of linear regression that applies a penalty (alpha) to the model coefficients in the regression cost function. Increasing alpha drives more coefficients toward zero, encouraging sparse solutions. To find the optimal penalty that balanced error due to overfitting and error due to bias in our models, we first used cross-validated lasso model fitting with an 80-20 train-test split. After identifying the penalty that minimized the mean squared error, we fit the lasso model a final time for each motor scale, using the optimized alpha. From this model, we identified the top 3 predictors with the largest magnitude coefficients, denoting the largest contribution to the model. **Step 4) Linear regression and bootstrapping**: We fit a linear regression model for each motor scale using the top 3 coefficients (limited to 3 to prevent overfitting) from lasso regression and adjusting for known predictors and confounders. Given their established association with motor outcome, global brain abnormality score on structural MRI^36^, male sex, gestational age at birth, PMA at MRI scan, and birth hospital were retained in all final models irrespective of their significance, to ensure the independent predictive power of our morphometric biomarkers. We verified that the final models met all assumptions of linear regression, including no multicollinearity, normality, additivity, no autocorrelation, and homoscedasticity. We used bootstrap resampling with 10,000 repetitions to ensure that the confidence intervals for our final models were stable.

### 2.6 Data and code availability statement

Code used in this analysis and derived data that support the conclusions of this study are available upon direct request to the corresponding author.

## 3. Results

### 3.1 Subjects

Of the initial 110 eligible VPT infants, 17 were excluded due to structural brain injury or issues with their T2 MRI scan: ten exhibited ventriculomegaly, one exhibited posterior plagiocephaly (flat head syndrome); which interfered with dHCP volume estimation; five had significant artifacts or missing cortical boundaries on their T2 images, and one was missing a T2 image altogether. Of the remaining 93 infants, 75 (81%) returned for follow-up testing at a mean (SD) age of 24.7 (2.8) months corrected age and were included in the analysis. Seven infants (9.3%) had moderate injury on structural MRI based on their global brain abnormality scores^35^. The mean (SD) Bayley-III motor score for the final cohort was 94.7 (13.2), with a range from 46 to122. Twelve infants (16%) scored more than 1 SD (≤85) below the mean. The mean (SD) raw fine and gross motor scores were 9.8 (2.5) and 8.3 (2.2), respectively. Additional baseline characteristics for the final cohort and for the infants lost to follow up are shown in Table 1. The two cohorts did not differ significantly on any baseline characteristics.

**Table 1.** Baseline characteristics of the final very preterm cohort with follow-up data included in the Bayley-III motor regression model (n=75).

| Characteristics | Final Cohort<br>with Bayley<br>Motor Follow-up<br>(N=75) | Lost to Follow-Up (N=18) | p |
| --- | --- | --- | --- |
| Chorioamnionitis, n (%) | 10 (13.3) | 4 (22.2) | 0.461 |
| Antenatal steroids,<br>complete course, n (%)* | 40 (54.8%) | 6 (33.3%) | 0.121 |
| Gestational age, weeks,<br>mean (SD) | 28.1 (2.4) | 28.6 (3.0) | 0.150 |
| Birth weight, grams,<br>mean (SD) | 1105.7 (370.1) | 1120.8 (385.1) | 0.438 |
| Male, n (%) | 44 (58.7) | 9 (50.0) | 0.599 |
| Severe retinopathy of<br>prematurity, n (%) | 8 (10.7) | 1 (5.6) | 1.000 |
| Bronchopulmonary<br>dysplasia, n (%) | 38 (50.7) | 9 (50.0) | 1.000 |
| Late-onset sepsis, n (%) | 7 (9.3) | 5 (27.8) | 0.051 |
| Postnatal steroids, n (%) | 6 (8.0) | 0 (0.0) | 0.592 |
| Low socioeconomic<br>status, n (%) | 41 (54.7) | 14 (77.8) | 0.109 |
| Global brain<br>abnormality score on<br>structural MRI, mean<br>(SD) | 3.2 (2.7) | 3.1 (3.5) | 0.264 |

\* Two infants in the final cohort did not have antenatal steroid information.

### 3.2 Cortical morphometrics

Total brain surface area was positively correlated with each Bayley motor scale, with PMA-corrected correlation coefficients of 0.52 for overall motor, 0.45 for gross motor, and 0.49 for fine motor. Total brain curvature was negatively correlated with each Bayley motor scale: -0.37 for overall motor, -0.34 for fine motor, and -0.33 for gross motor. Total brain sulcal depth was positively correlated with overall motor score (0.29) and gross motor score (0.28), but not with fine motor score. Total brain gyrification index was also significantly correlated with overall motor score (0.27) and gross motor score (0.29), but not with fine motor score.

Figure 1 illustrates the strength of the correlation between each *lobar* cortical morphometric and all three scales, adjusted for PMA at MRI scan. Cortical surface area was positively correlated with every motor scale for every lobe of the preterm brain, with correlation coefficients ranging from 0.30 to 0.57. Cortical curvature was negatively correlated with each motor scale in several lobes of the brain, with correlation coefficients from -0.26 to -0.43. For overall motor score, correlations existed in the bilateral frontal and parietal lobes, the left temporal lobe, and the left insula. For fine motor score, correlations existed in the left frontal lobe, the left insula, the left temporal lobe, and the right parietal lobe. For gross motor score, correlations existed in the bilateral frontal and parietal lobes, the left insula, and the left temporal lobe. Gyrification index was correlated with overall motor score in the right occipital lobe and with gross motor score in the left parietal lobe and the right occipital lobe, with correlation coefficients from 0.26 to 0.29. Sulcal depth was correlated with overall motor score in the right parietal lobe and with gross motor score in the left frontal lobe, with correlation coefficients from 0.26 to 0.28.

**Figure 1:**
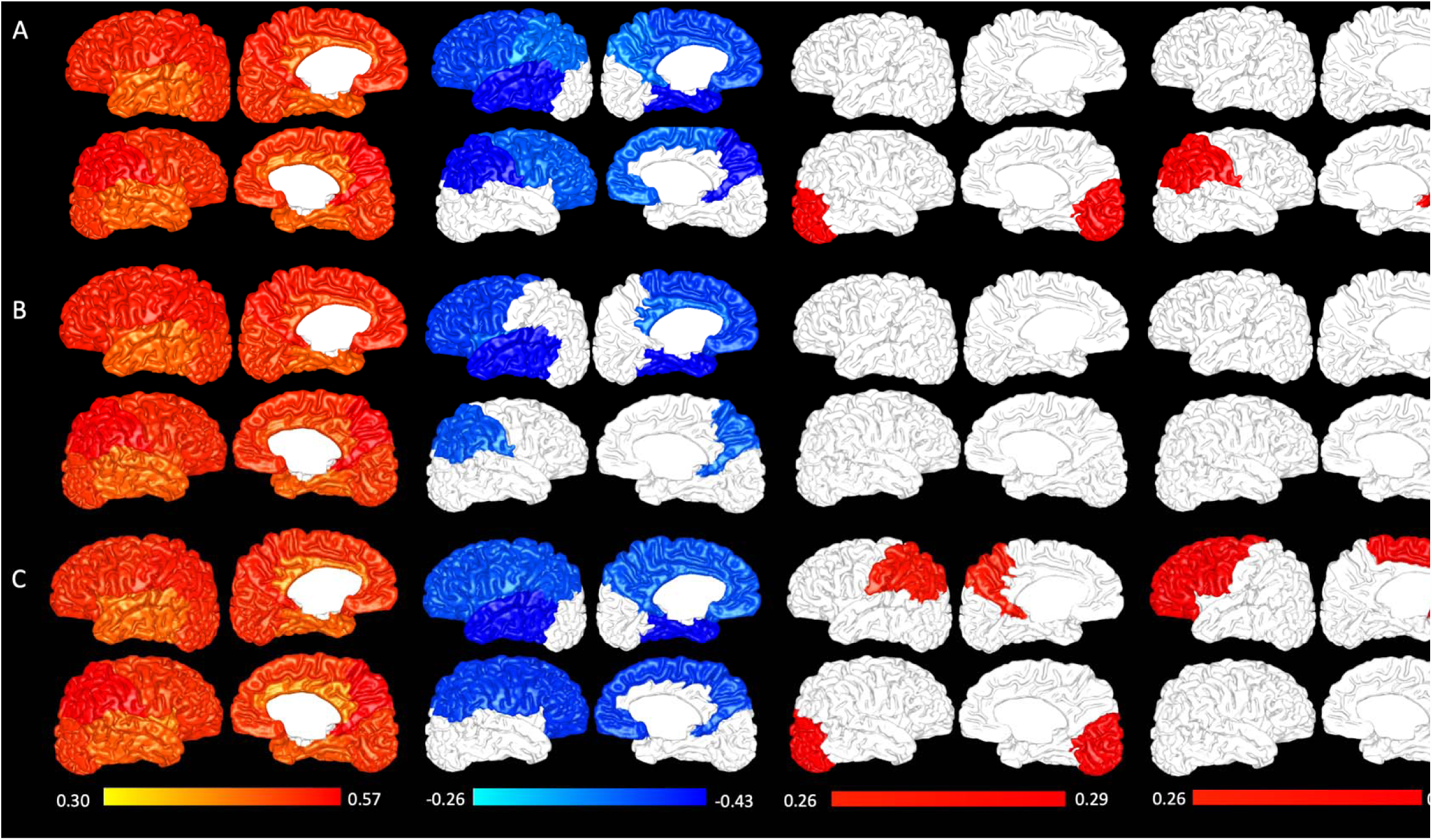
Whole-lobe partial correlations (PMA-adjusted) between cortical morphometrics and Bayley-III motor outcomes. Row A displays correlations between Bayley-III overall motor score and cortical surface area (column 1), inner cortical curvature (column 2), gyrification index (column 3), and sulcal depth (column 4). Rows B and C display the same correlations for the Bayley-III fine and gross motor subscale scores, respectively.

### 3.3 Volumetrics

Total brain volume was positively correlated with each motor scale: 0.43 for overall motor, 0.37 for gross motor, and 0.41 for fine motor. In terms of overall tissue volumes, white matter volume (0.37), cortical grey matter volume (0.42), and deep grey matter volume (0.52) were all positively correlated with overall motor score. White matter volume (0.33), cortical grey matter volume (0.35), and deep grey matter volume (0.50) were positively correlated with gross motor score. White matter volume (0.34) and deep grey matter volume (0.46) only were positively correlated with fine motor score. Volume of the ventricles and volume of overall CSF were not significantly correlated with any motor scale.

Figure 2 illustrates the correlation strength between each lobar and subcortical volume and the Bayley-III motor scales, adjusted for PMA at MRI scan. All five bilateral lobar volumes were positively correlated with all motor scales, with the exception of the temporal lobe bilaterally for the gross motor scale. Lobar volume correlations ranged from 0.29 to 0.47 for overall motor,0.33 to 0.41 for gross motor, and 0.28 to 0.45 for fine motor. Nearly all subcortical volumes tested were positively correlated with all motor scales. Overall motor score was positively correlated with volume: bilaterally in the amygdala, the caudate, the lentiform nucleus, and the subthalamic nucleus; in the left hippocampus; and in the brainstem, the cerebellum, the corpus collosum, and the thalamus, with correlations ranging from 0.29 to 0.55. Gross motor score was positively correlated with volumes: bilaterally in the caudate, the lentiform nucleus, and the subthalamic nucleus; in the right amygdala; and in the brainstem, cerebellum, and thalamus, with correlations ranging from 0.32 to 0.52. Fine motor score was positively correlated with volume: bilaterally in the amygdala, the lentiform nucleus, and the subthalamic nucleus; in the left caudate and the left hippocampus; and in the brainstem, cerebellum, corpus callosum, and thalamus, with correlations ranging from 0.27 to 0.50. Thalamic volume was the most highly-correlated subcortical volume for all motor scales.

**Figure 2:**
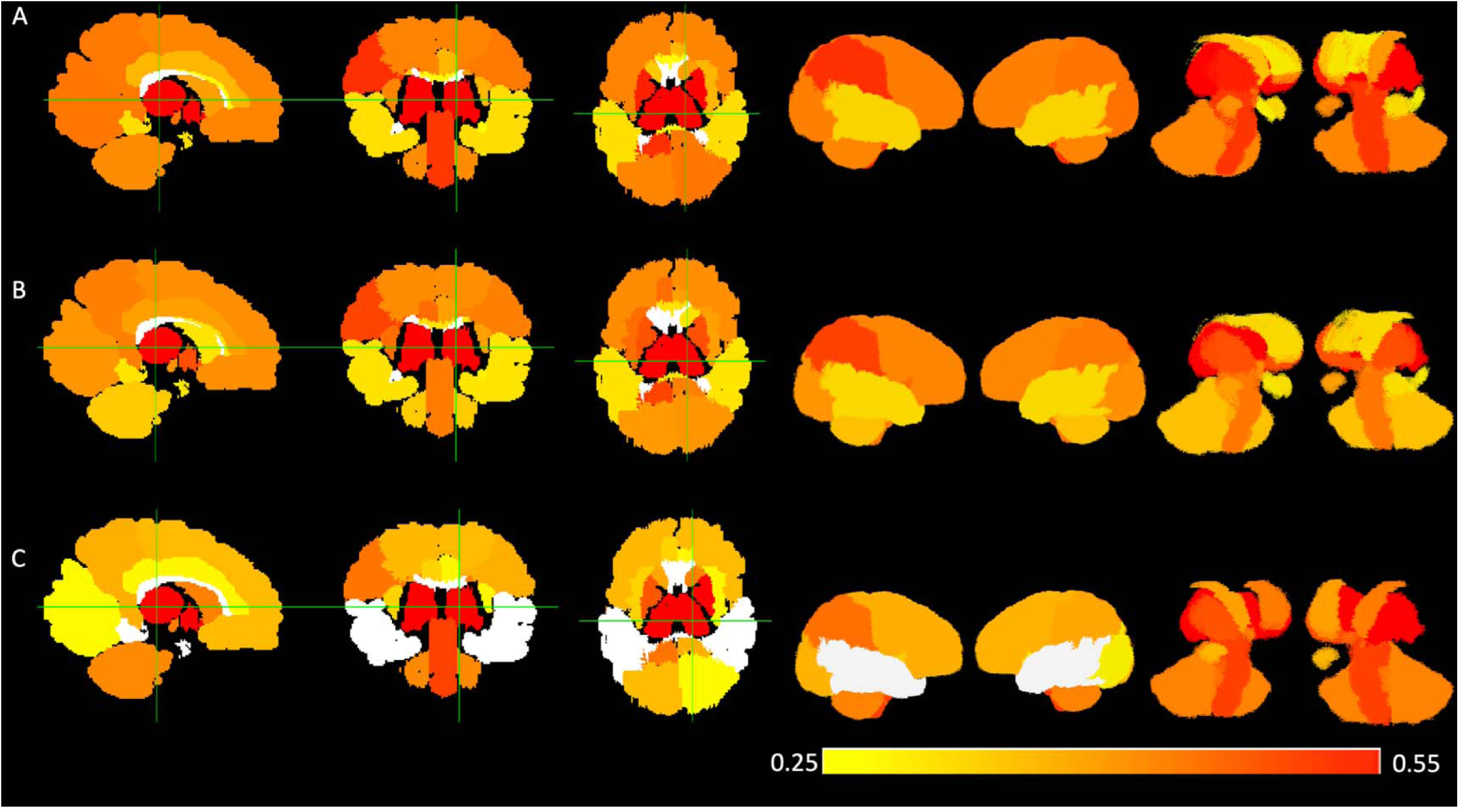
Partial correlations (PMA-adjusted) between brain volumes and Bayley-III motor outcomes. From left to right, row A displays correlations between Bayley-III overall motor score and 1) all brain volumes shown in 2-D from sagittal, coronal, and axial perspectives, 2) all brain volumes shown in 3-D from right and left perspectives, and 3) subcortical structures only shown in 3-D from right and left perspectives. Rows B and C display the same correlations for the Bayley-III fine motor subscale score and the Bayley-III gross motor subscale score, respectively.

### 3.4 Regression results

Our automated feature-selection method using lasso regression with an optimized penalty selected the following best three predictors for the overall motor scale: thalamic volume, curvature of the left temporal lobe, and curvature of the left insula. For the fine motor scale, the top three predictors were: surface area of the right occipital lobe, curvature of the left temporal lobe, and curvature of the left insula. For the gross motor scale, the top three predictors were: thalamic volume, curvature of the left temporal lobe, and gyrification index of the parietal lobe. Table 2 displays the final regression models for all three Bayley-III motor scales, with and without known predictors added. All cortical morphometrics and volumetrics remained significantly correlated with motor outcomes when known predictors were introduced into the models, illustrating the independent predictive power of these biomarkers. For each motor scale, the combination of just three automatically-extracted and automatically-selected cortical morphometric and volumetric features explained over one-third of the variance in motor scores. In the case of overall motor score, the three features explained nearly one-half of the variance.Global brain abnormality score on structural MRI and PMA at MRI scan were both negatively correlated with the overall and gross motor scales. The negative correlation with PMA at MRI scan is likely due to the sicker babies in our cohort being scanned at slightly later time points, when they were healthy enough to safely undergo MRI. Male sex was negatively correlated with gross motor score. The bootstrap bias-corrected confidence intervals for all three final models were comparable (Table 2), which supports their internal validity.

**Table 2.**
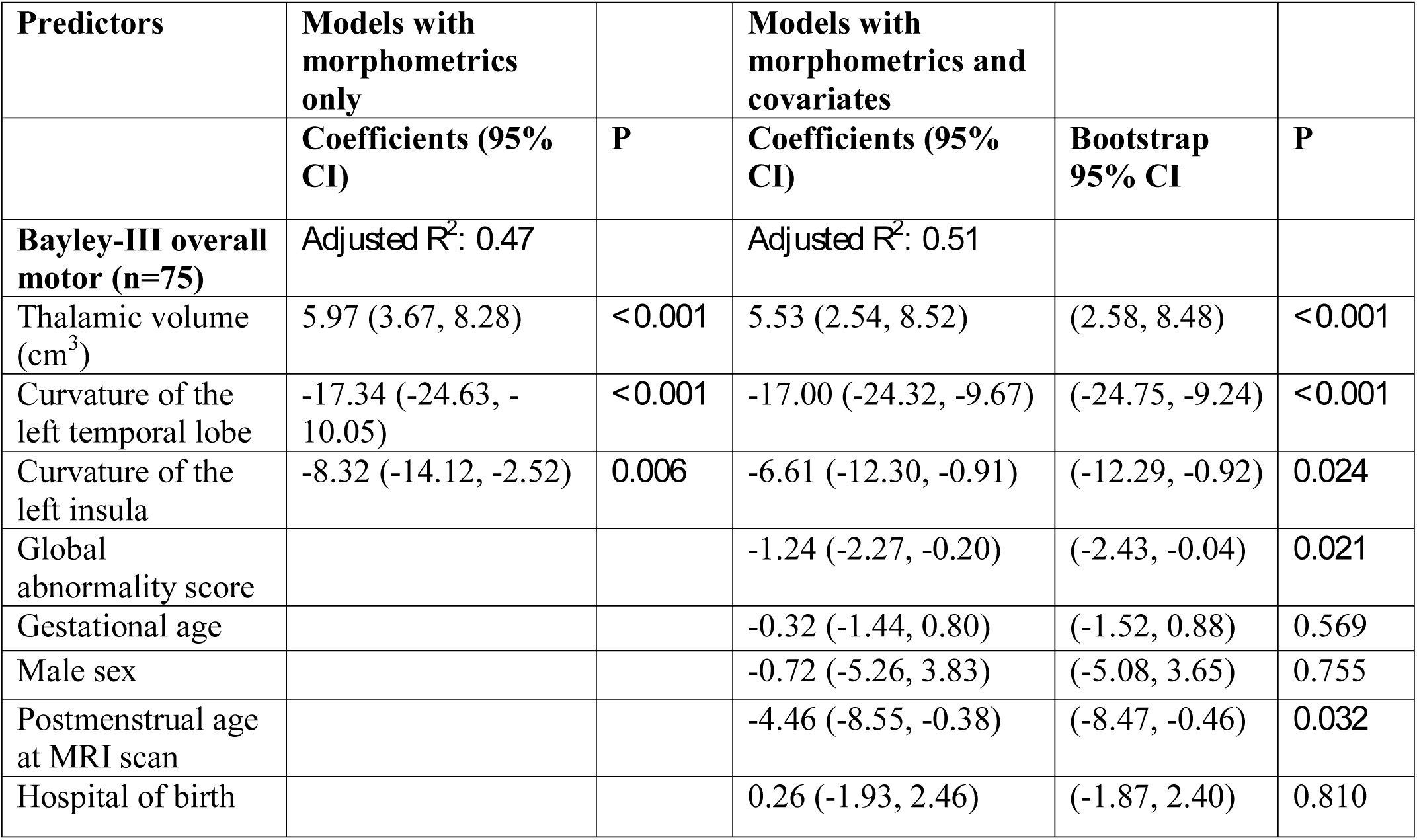
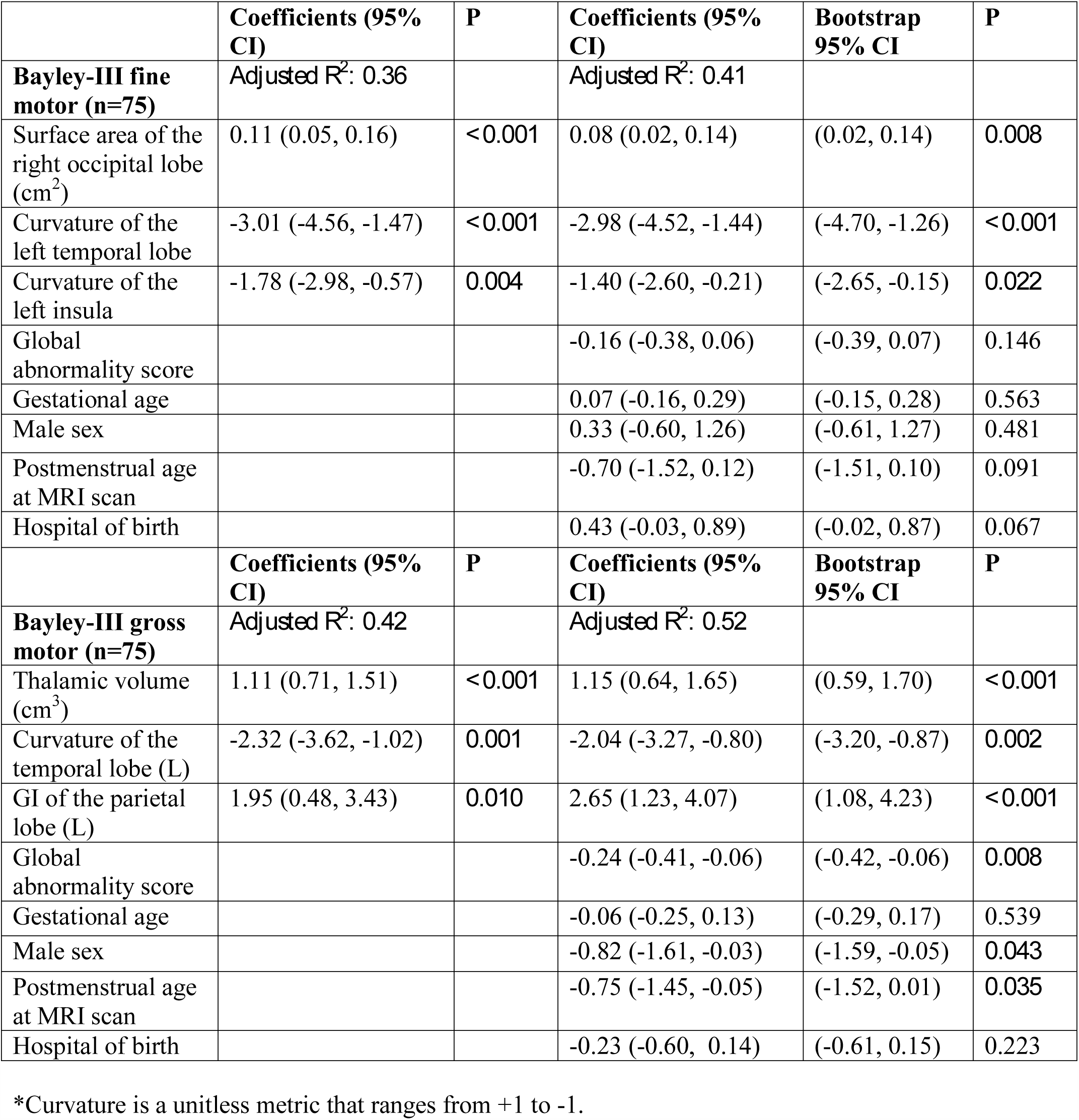
Morphometric biomarkers and prediction of **1)** Bayley-III motor scores with and without adjustment for known predictors and confounders (global injury score on structural MRI, gestational age, and male sex).

## 4. Discussion

In this study, we illustrate that motor ability at two-years corrected age in a preterm cohort can be independently predicted with brain morphometrics and volumetrics automatically extracted from structural MRI at term. This is especially true for thalamic volume, cortical surface area,and cortical curvature. Greater global and lobar cortical surface area, greater overall volume of the whole brain and of most tissue types, and greater subcortical volumes were positively correlated with all scales of two-year motor ability. Greater cortical curvature was negatively correlated with all scales, both globally and in the majority of lobes. Greater gyrification and sulcal depth were positively correlated with overall motor score and gross motor score, globally and in one to two brain lobes each. In multivariable regression, just three automatically-extracted and automatically-selected cortical morphometric and/or volumetric biomarkers accounted for between a third and a half of the variance in fine, gross, and composite scales of motor development at 2 years corrected age.

Inner cortical curvature is a novel prognostic biomarker for motor ability. Makropoulos et al. previously reported increased inner cortical curvature throughout the preterm brain as compared to term controls^22^, and we corroborated this finding in our very preterm cohort^23^. Curvature of the left insula and curvature of the left temporal lobe were both retained in overall motor and fine motor models. Curvature is likely such a valuable metric for prediction because it generalizes some of the major brain maturational abnormalities associated with prematurity. Premature infants consistently exhibit shallower sulci and decreased gyrification compared to their full-term peers. Shallower sulci are missing sulcal wall area (low curvature), but have the same amount of sulcal trough area (high curvature), which results in higher average curvature^29^, although the individual anatomy of sulci and gyri can vary considerably. The insula is involved in some aspects of motor and somatosensory control^37^ while regions of the temporal lobe help to mediate visuomotor coordination^38^. Taken together, the curvature of the insula and the curvature of the temporal lobe explained 28% of the variance in the overall motor model and 22% of the variance in the gross motor model.

The association between brain volumes and motor ability in preterm cohorts has been well-established. Some studies have identified this relationship at the same developmental timepoint, rather than using volumes to prognosticate later motor outcomes^16,39^. At seven years, basal ganglia and thalamic volumes were associated with motor function^39^, and at three years, volumes of the left and right sensorimotor cortices were associated with gross motor score^16^. Still other prognostic studies, have examined associations between only a few select brain volumes at TEA and motor outcome; for instance reduced cerebellar volume and abnormal general movements at three months^15^ or unmyelinated white matter volume and two-year motor outcomes^17^. Gui et al. showed that a classifier trained using brain volumes from TEA was better able to predict psychomotor developmental index score at 18–24 months^13^ than a classifier trained on social and demographic variables alone, but they did not examine which brain volumes were the most predictive. Specific brain volumes from TEA have been shown to prognosticate motor ability into late childhood. Volumes of the cerebellum and brainstem predicted fine motor skills at age six^19^, and volumes of the frontal lobes, basal ganglia, thalamus, cerebellum, and brainstem were correlated with motor scores at age 11^18^. Our results corroborate and extend prior studies by showing positive correlation between numerous subcortical volumes at TEA — the volume of the amygdala, brainstem, caudate, cerebellum, hippocampus, lentiform nuclei, subthalami nuclei, and thalamus—and motor outcomes at two-years corrected age. Like Loh et al.^39^, we identified the thalamus as a top predictive volumetric biomarkers for motor development. Thalamic volume was a top-three predictive biomarkers for the overall motor and gross motor ability. In bivariable regression, thalamic volume alone accounted for 27% of the variance in overall motor score and 26% of the variance in gross motor score. The thalamus is a crucial motor hub that relays signals from the cerebellum and basal ganglia to the cerebral cortex, which may explain why it is such a prominent motor biomarker.

We found that surface area of the very preterm brain was positively correlated with all Bayley-III motor scales, globally and bilaterally in all five lobes, making it the most universally-correlated metric tested. Comparing our results with those in the literature, the robust relationship we identified between cortical surface area and motor ability, appears to persist into childhood and early adulthood. Grunewalt et al. identified reduced cortical surface area and poorer motor skills in a cohort of extremely-low-birth-weight children at age ten^9^. In preterm-born young adults (18-22 years of age), Sripada et al. demonstrated a positive relationship between measures of visual motor integration and cortical surface area in all lobes, with associations in the temporal lobe, the insula, and the occipital lobe being the most consistently significant^32^. In the present study, the surface area of the right occipital lobe explained 18% of the variance in the fine motor subscale, presumably reflecting the high level of visual input and feedback needed to perform a fine motor task. The fact that surface area was not retained in the overall or gross motor models was likely due to the high degree of collinearity between thalamic volume and surface area. In our final cohort, overall surface area was significantly correlated with thalamic volume (r=0.79, p≤0.0001). Conversely, overall curvature was not (r= -0.10, p= 0.41), potentially explaining why curvature biomarkers were retained in the final models with thalamic volume.

Our study was limited by a modest sample size and 19% loss to follow-up for our primary analysis. However, the infants lost to follow-up did not differ from included infants on any baseline characteristics, which should minimize attrition bias. Given that only seven subjects in our final cohort developed cerebral palsy, we lacked sufficient study power to examine prediction of cerebral palsy as an important outcome.

## 5. Conclusions

In conclusion, of the numerous biomarkers available on structural MRI at TEA, thalamic volume and/or whole lobar surface area combined with whole lobar measures of curvature appear to be the best prognostic biomarkers for early motor development in very preterm infants. Gyrification index of the left parietal lobe also improves prediction of gross motor ability. Using just three automatically extracted metrics, we were able to account for nearly one-half of the variance in overall Bayley-III motor score at two-years corrected age. Overall, cortical morphometrics and volumetrics automatically-derived from structural MRI at term show promise to prognosticate later motor ability in very preterm infants. Larger cohort studies are needed to validate these biomarkers and to develop additional advanced MRI biomarkers to enhance the models’ predictive power and facilitate aggressive early intervention for the most at-risk infants.

## Data Availability

Derived data supporting the conclusions of this study are available upon request to the corresponding author.

## 6. Author contributions

**Julia E. Kline**: Data Curation, Formal analysis, Writing - original draft, Writing - review & editing, Investigation, Methodology, Resources, Software, Validation, Visualization. **Venkata Sita Priyanka Illapani:** Resources, Writing - review & editing, Validation. **Lili He:** Data curation, Writing - review & editing. **Nehal A. Parikh:** Conceptualization, Data curation, Formal analysis, Writing - review & editing, Funding acquisition, Investigation, Methodology, Project administration, Resources, Supervision.

## 7. Acknowledgements

We thank Jennifer Notestine and Valerie Marburger for study coordination, Josh Goldberg for recruitment assistance, Mark Smith for serving as our lead MR technologist, and the families and NICU personnel that made this study possible. National Institutes of Health grants R01-NS094200 and R01-NS096037 from the National Institutes of Neurological Disorders and Stroke and R21-HD094085 from the Eunice Kennedy Shriver National Institute of Child Health & Human Development supported this study. The funders had no role in study design, data collection and analysis, and writing or submission of this manuscript.

## Abbreviations

VPT: Very preterm
GA: gestational age
TEA: term-equivalent age
MRI: 3magnetic resonance imaging
CP: cerebral palsy
PMA: postmenstrual age
dHCP: developing Human Connectome Project
SD: standard deviation

